# Awake Prone Position in Hypoxemic Patients with Coronavirus Disease 19 (COVI-PRONE): A Study protocol and Statistical Analysis Plan for Randomized Clinical Trial

**DOI:** 10.1101/2021.08.06.21261531

**Authors:** Zainab Al Duhailib, Yaseen M Arabi, Sarah Culgin, Jason Weatherald, Ken Kuljit S. Parhar, Kate Nelson, Hani Tamim, Waleed Alhazzani, the COVI-PRONE Trial investigators

**Author notes:** **Correspondence:** Zainab Al Duhailib, MBBS, EDIC, MSc (epid), King Faisal Specialist Hospital and Research Centre, Riyadh, Saudi Arabia.

## Abstract

**Background:** Coronavirus disease 2019 (COVID-19), may progress to respiratory failure requiring invasive mechanical ventilation. Due to ventilator shortage and healthcare systems strain, affordable interventions such as awake prone positioning has been used to improve oxygenation, however, the effect of this intervention on patient-important outcomes is uncertain. The COVI-PRONE trial aims to determine if awake prone positioning in hypoxemic COVID-19 patients reduces the need for invasive mechanical ventilation.

**Study design:** A pragmatic, multicenter, international, parallel-group, and stratified randomized controlled trial, aiming to enrol 400 hospitalized adults with COVID-19.

**Participants:** The target population is hospitalized adults with confirmed or suspected COVID-19, hypoxemia that requires ≥40% oxygen or ≥ 5 L/min by nasal cannula, and abnormal chest x-ray. We will exclude patients with any of the following: immediate need for intubation; altered mental status; contraindication to prone positioning; hemodynamic instability; body mass index > 40 kg/m^2^; third trimester pregnancy; do not intubate status; previous enrolment or intubation within the same hospital admission; and prone positioning for more than one day prior to randomization.

**Study intervention and control:** Following informed a priori or deferred consent, eligible patients will be centrally randomized to either the intervention arm (prone positioning) or standard of care (no prone positioning). Patients randomized to the prone position will be required to either self-prone or assist-prone for a total of eight to ten hours per day until they meet pre-specified stopping criteria.

**Study outcomes:** The primary outcome is invasive mechanical ventilation at 30-days of randomization. Other outcomes include mortality at 60 days, invasive and non-invasive mechanical ventilation free days at 30 days, hospital length of stay at 60 days, days alive and outside of the hospital at 60 days, complications of proning, and serious adverse events.

## 1.0 Background

Since early 2020, the world has struggled with the coronavirus disease 2019 (COVID-19) pandemic (1). COVID-19 causes acute respiratory distress syndrome (ARDS) in up to a third of hospitalized patients (2). This translates into thousands of patients requiring admission to the intensive care unit (ICU) (3, 4).

The search for effective and affordable interventions continued as the pandemic unfolded. Awake prone positioning is a novel and potentially attractive therapeutic intervention. Proning was first used in 1976 for five patients with ARDS and refractory hypoxemia, which resulted in improved oxygenation and enhanced drainage of pulmonary secretions (5). Since then, evidence has evolved, and prone positioning is currently used in invasively ventilated patients with moderate-severe ARDS.

Recent clinical practice guidelines also recommend prone positioning in mechanically ventilated patients with moderate-to-severe ARDS (6), including patients with COVID-19 (7). Despite the rich literature supporting prone positioning in this population, there are no large multicentre randomized trials examining the effect of awake prone positioning in non-intubated patients.

A rapid review identified 35 observational studies (n= 414 patients) on awake prone positioning in patients with hypoxemic respiratory failure including those with COVD-19 (8). Almost all studies showed a transient improvement in oxygenation with prone positioning. Subsequently, a few small randomized controlled trials (RCTs) were published on this topic (9-11).

Given the ventilator shortage and the strain of the healthcare system during the recurrent waves of the pandemic, interventions that could prevent endotracheal intubation would help to conserve scarce resources and improve patient-outcomes. Therefore, we planned a multicentre RCT to determine the efficacy and safety of awake prone positioning in non-intubated patients with COVID-19 and hypoxemic respiratory failure. If proven effective, this intervention could be widely adopted even in low-income countries.

## 2.0 Study design and methods

The *Awake Prone Position in Hypoxemic Patients with Coronavirus Disease-19* (COVI-PRONE) trial is designed as a multicentre, parallel-group, stratified randomised controlled trial (RCT) that will take place at centres in Canada, Saudi Arabia, Kuwait, and the USA. The trial was prospectively registered in *ClinicalTrials*.*gov* (NCT04350723) (12). The design and conduct of the trial will follow the SPIRIT guideline (13), and will be reported according to the CONSORT Statement (14).

### 2.1 Objective

To determine the effect of awake prone positioning (versus no prone positioning) in non-intubated hypoxemic COVID-19 patients on the need for invasive mechanical ventilation.

### 2.2 Hypothesis

We hypothesize that awake prone positioning of non-intubated hypoxemic COVID-19 patients will reduce the need for invasive mechanical ventilation compared to no proning.

### 2.3 Study population

Eligible patients should meet all the following criteria: adults ≥ 18 years of age; confirmed or suspected COVID-19; hypoxemia that requires ≥40% oxygen or ≥ 5 L/min by nasal cannula; bilateral or unilateral chest infiltrates on chest x-ray; and admission to the ICU or an acute care unit where hemodynamic and respiratory monitoring is available.

We will exclude patients with any of the following: immediate need for intubation; decreased level of consciousness (Glasgow Coma Scale score <10) or significant cognitive impairment that may interfere with compliance (i.e. delirium, dementia); presence of any contraindication to prone positioning, significant bony fractures or abnormalities that interfere with prone positioning; complete bowel obstruction; active upper gastrointestinal bleeding, patient is unable to prone, or likely will not comply with proning; hemodynamic instability; body mass index (BMI) > 40 kg/m^2^; pregnancy in the third trimester; patient or substitute decision maker (SDM) or caring physician refuses enrolment in the study; do not intubate status; previous enrolment and intubation within the same hospital admission; and prone positioning for more than one day prior to randomization. Details of inclusion and exclusion criteria are presented in **Table 1**.

**Table 1:**
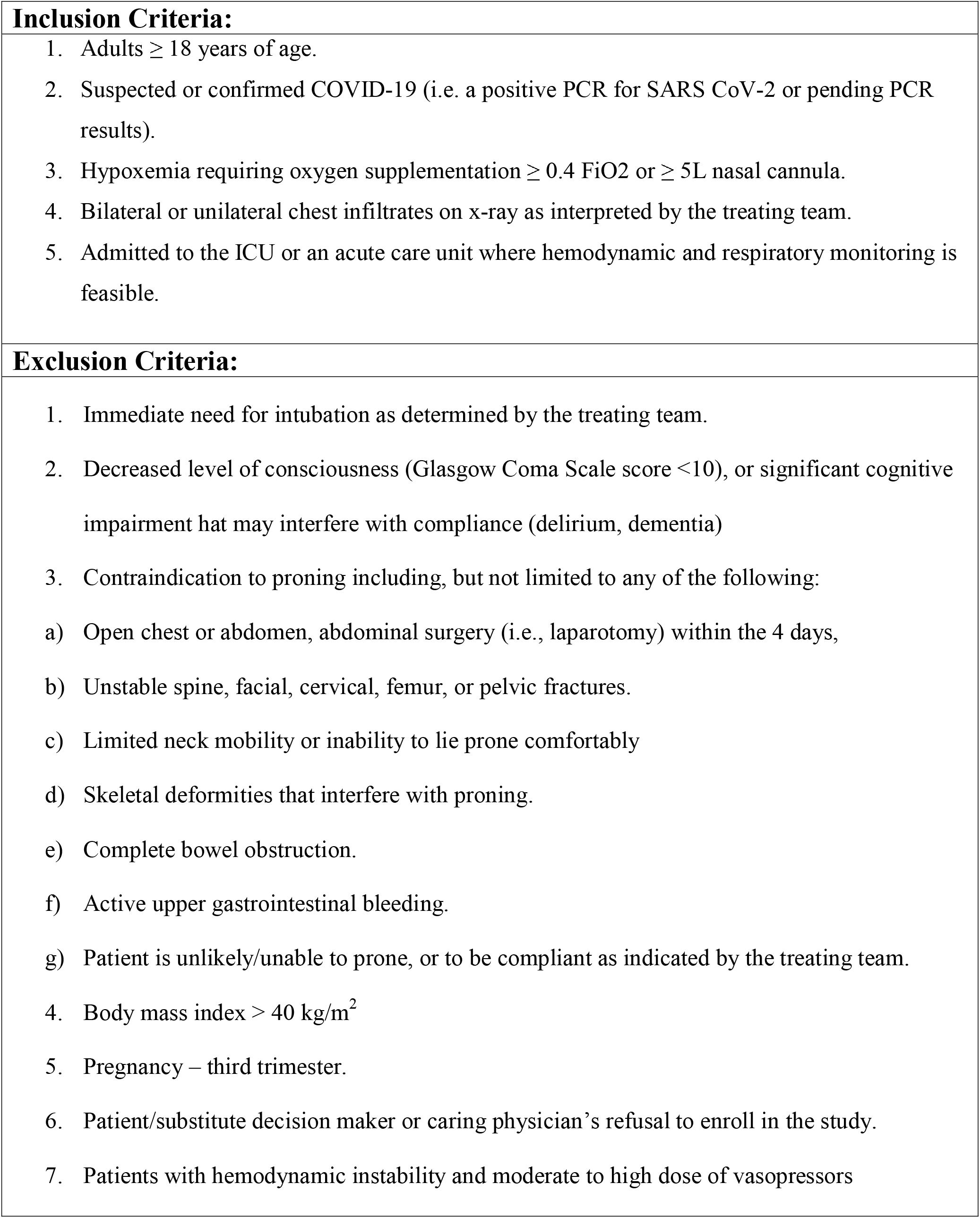
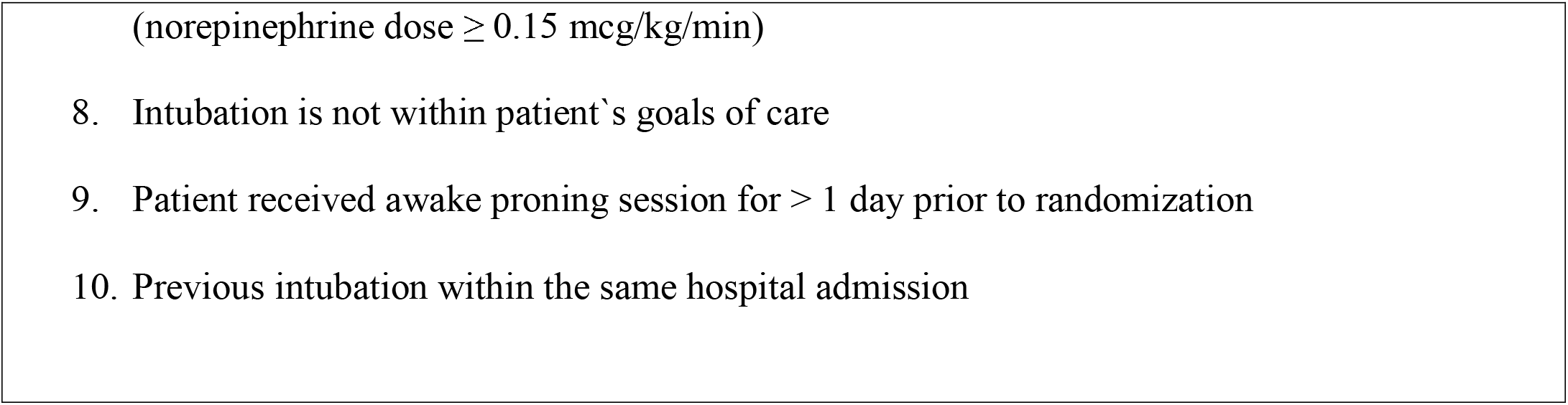
Eligibility Criteria.

### 2.3 Consenting model

We will obtain research ethics board (REB) approval at each participating centre. We plan to enrol patients who are acutely ill and in respiratory distress, which will hinder their ability to provide first-person informed consent. As such, we anticipate that many eligible patients will be randomized using a deferred consent model. After eligible patients have been identified, the research team will make every effort to contact the SDM, if one has already been identified and is able to be contacted by phone. Successful contact with the SDM may not always be the case during the period of the patient’s eligibility-especially with the applied policy on visitor restriction to the hospital to limit transmission. Since our study aims to initiate proning as soon as eligibility criteria have been met, and to avoid rapid deterioration and further lung injury, rapid enrolment is crucial. If the SDM is not able to be contacted within 15 minutes of eligibility, the eligible patient will be enrolled and randomized. Consent will be ongoing, and full informed consent will be obtained from the patient or their SDM when they are available and able to provide informed consent. Should the SDM or the patient refuses to continue to participate in the study, this request will be honoured but data obtained to this point will be retained and permission will be sought for continued data collection. For participants whom no SDM can be located, if the patient dies or does not recover decision-making capacity, we will retain the data as per local REB procedures.

In extenuating circumstances, patients may be able to adequately provide *a priori* informed consent. *A priori* consent will be sought if GCS is 15 out of 15 and a suitable method to communicate with the patient within the isolation environment is available.

### 2.4 Randomization

The research team will screen all patients aged ≥18 years who are admitted to the ICU or acute care unit for eligibility. Randomization of eligible study participants will be at 1:1 ratio and based on variable undisclosed block sizes (two, four, and six blocks) through a central computer-generated table using Randmoize.Net website (http://www.randomize.net/). The randomization algorithm prepared by a biostatistician will stratify patients by the site of recruitment and severity of hypoxemia. The hypoxemia severity will be classified based on the ratio of peripheral oxygen saturation to fractional inspired oxygen (SpO_2_:FiO_2_) into two strata: SpO_2_:FiO_2_ > 150 and SpO_2_:FiO_2_ ≤ 150 immediately prior to randomization. We estimated the SpO_2_:FiO_2_ thresholds using the following equation: SpO_2_/FiO_2_ ratio = 68 + 0.84 x (PaO_2_/FiO_2_ ratio) (15). Study participants will be randomized to either the intervention arm (i.e., prone positioning) or the control arm (i.e., no prone positioning).

### 2.5 Allocation Concealment

Concealment of randomization will be ensured at each centre through a remote dedicated online randomization system. Post randomization blinding is not possible due to the nature of the prone positioning procedure.

### 2.6 Study Procedure

The Research Coordinator at each centre will screen new admissions to the ICU or the acute care unit daily for patients with suspected or confirmed COVID-19. Patients fulfilling inclusion criteria without meeting any exclusion criteria will be eligible for enrolment.

#### 2.6.1 Intervention arm

In addition to receiving usual care, patients randomized to the intervention arm will be in prone positioning for 8-10 hours daily. Patients will be allowed 2 to 3 breaks (1-2 hours each) in supine position, if needed. Prone positioning will continue daily until one of the following stopping criteria is met: 1) a relative improvement in FiO_2_ requirement from baseline by 40% that is sustained for 24 hours, 2) the patient is intubated, or 3) the patient is discharged from the ICU or acute care unit.

Patients who can self-prone will do so under supervision of the treating team. For patients who are unable to self-prone or require assistance; the treating team will assist the patient in proning. Then, place the cardiac leads on the right and left shoulders of the patient along with one on the left pelvis, intravenous access and tubes will be secured in one side. The healthcare team may consider placing three pillows under the patient to ensure comfort. One pillow will be placed at the head of the bed, the second will be placed below the chest, and the third under the knees. The patient will be moved away from the side of oxygen tubing and lines with the assistance of two healthcare workers, which is away from the side the patient will be turned to. Then, the patient will be turned to their side. The patient will be assisted to turn his body with head facing the bed. This will be followed by lifting one arm to the side that the patient is facing for comfort. Once the patient is in the prone position, the head will be adjusted to face the side of the device (videos attached, https://cutt.ly/Ayk1iZt). The patient’s vital signs (heart rate [HR], mean arterial pressure [MAP], and SpO_2_) will be monitored as per the institution’s protocol.

Failure of prone positioning will be defined as: endotracheal intubation, increasing FiO_2_ by 20% from baseline after 2 hours from initiation of proning, or persistent discomfort or agitation that is not controlled by pharmacologic agents.

#### 2.6.2 Control arm

Patients randomized to the control arm will receive usual care at the discretion of the treating team but without prone positioning.

The choice of modality of oxygen delivery in the intervention or the control arm (i.e., nasal cannula, face mask, HFNC, or NIPPV) will be at the discretion of the treating team. In both arms, the patient will be observed for 15 minutes to ensure that they are tolerating the current oxygen delivery device.

#### 2.6.3 Protocol Deviation

Protocol deviation in the intervention arm is defined as zero hours of proning across all days prior to meeting any of the intervention stopping criteria. While protocol deviation in the control arm is defined as the any number of hours of proning across all days up to meeting stopping criteria.

### 2.7 Outcomes

- The **primary outcome** is invasive mechanical ventilation within 30 days of randomization. **Secondary outcomes** include: 1) mortality at 60 days, 2) Invasive mechanical ventilation free days at 30 days (defined as days alive and free of invasive or mechanical ventilation), 3) Non-Invasive ventilation free days (defined as days alive and free of non-invasive ventilation) 4) Hospital length of stay truncated at 60 days (defined as days alive and outside of ICU [i.e., ICU free days]), 4) Days alive and outside of the hospital truncated at 60 days (defined as days alive and outside of hospital), 5) Complications of proning (defined as accidental removal of intravenous access, hypotension, falls, pressure ulcers, or other adverse events. **Safety outcomes** include serious adverse events (any adverse event occurring following study mandated procedures, directly related to the treatment or intervention that results in any of the following outcomes: death, life-threatening adverse event, disability, or incapacity).

## 3.0 Statistical Analysis

### 3.1 Sample Size Calculation

We originally estimated the study sample size to be 350 patients. This was based on a relative risk reduction (RRR) of 25% (15), a 60% intubation rate in the control arm (16), and a power of 80%, and two-sided type I error of 5%. As the management of COVID-19 evolved, the threshold for intubation of COVID-19 patient changed, recent guidelines suggest a trial of HFNO or NIPPV prior to intubation. Therefore, we increased the sample size by 14% (i.e., target sample size of 400 participants) which allows 80% power to detect 30% RRR in the primary outcome given a control group event rate of 45%. This change was approved by both the trial’s Steering Committee and the Data Safety and Monitoring Committee (DSMC).

### 3.2 Data analysis

The detailed statistical analysis plan (SAP) is provided in the **supplement**. We will present patients’ demographics and clinical characteristics as frequencies and percentages for categorical data and mean and standard deviation (SD) or median and interquartile range (IQR) for continuous data as appropriate.

Given the nature of the intervention, it is not possible to blind the trial biostatistician to the study group for the interim or final analyses, however, the investigators and other methods team members will remain blinded until analysis is of complete outcome data is finalized. We will use the intention-to-treat principle, in which trial participants will be analysed in the group to which they were randomized.

The primary analysis will be an unadjusted survival analysis (time-to-event) taking into account the two stratified randomized variables of severity of hypoxemia and centre. In this analysis we will use all information up to the time of censoring. We will calculate hazard ratios (HR) and associated 95% confidence intervals (CI) using Cox regression analysis.

For continuous outcomes (i.e., hospital length of stay and ventilator free days), we will compare the two arms using a nonparametric approach to compare median durations because the distributions are likely skewed. We will also compare survival distributions using a log-rank test for the time-to-event analyses. Continuous outcomes will be analyzed using multiple linear regression where our main independent variable will be randomized group, and we will also include recruitment site and severity of hypoxemia as covariates.

**The secondary analysis** will compare the proportion of patients in the two groups with the binary primary and secondary outcomes using the χ2 test or Fisher exact test. We will calculate the relative risks and 95% CI. If there is a clinically important and/or statistically significant difference between groups and if otherwise appropriate, we will calculate metrics such as the number needed to treat or number needed to harm.

#### Effectiveness analysis

We will conduct an effectiveness analysis (i.e., as treated analysis) that excludes participants who (a) were randomised in error, (b) withdrew consent early, (c) never underwent prone positioning in the intervention arm, or (d) crossed over to routine prone positioning from the control arm.

#### Efficacy analysis

We will also conduct an efficacy analysis including all patients who undergone at least 8 hours of prone positioning daily until meeting discontinuation criteria (in the intervention arm) and those who did not undergo any hours of prone positioning (in the control arm).

Complications will be presented per randomized group as frequencies and percentages. Statistical significance will be set at alpha = 0.05. All analyses will be performed using SAS version 9.4 (Cary, NC).

### 3.3 Subgroup Analysis

The details on subgroup analysis are provided in the **supplement**. We plan to conduct five subgroup analyses based on: 1) severity of hypoxemia at randomisation classifying subgroups into SPO2:FiO2 ratio > 150 versus ≤ 150 (we hypothesize that the treatment effect will be larger in patients with severe hypoxia); 2) disease status classifying subgroups into SARS CoV-2 positive patients versus patients with negative testing for SARS CoV-2 (we hypothesize that the treatment effect will be larger in the SARS CoV-2 positive subgroup); 3) Oxygen delivery mode: nasal cannula or oxygen mask versus NIPPV versus HFNO (we hypothesize that HFNO group will have a larger treatment effect); 4) age ≥70 or <70 years old (we hypothesize that younger patients will have a larger treatment effect); 5) sex.

All subgroup analyses will only be performed for endotracheal intubation outcome and will be summarized visually using Forest plots. We will use the test of interaction to compare subgroups. Results will be reported using HR and 95% CI and the Cox regression analysis will be used to report the results of tests of interactions.

We plan to conduct the following sensitivity analyses: 1) death as a competing risk; 2) per-protocol analysis; 3) per-site analysis. For details on sensitivity analyses, please refer to the SAP in the **supplement**.

## 4.0 Study Management

### 4.1 Data Safety and Monitoring Committee (DSMC)

An independent DSMC, that includes an experienced critical care trialist, academic critical care clinicians, and an experienced, independent, academic biostatistician, will review the accumulating study data after 50% of patients have been randomised and make recommendations to the Steering Committee about the conduct of the trial, integrity of the data and trial discontinuation to ensure the overall safety of participants. An independent biostatistician will conduct the interim analysis following the statistical analysis plan. The DSMC will use a modified Haybittle-Peto method for benefit or harm as a general guidance for decision making (17, 18). The DSMC will also be monitoring the safety of the participants, focusing on adverse events and other safety indicators. Serious adverse events will be reported to the DSMC chair on regular basis.

### 4.2 Data collection, storage, and monitoring Plans

#### 4.2.1 Data Collection

Research Coordinators will enter the study data in a structured Data Collection Form using REDCap online software (www.project-redcap.org). The following variables will be recorded or calculated: demographic data (age, sex, body mass index [BMI] and co-morbidities); severity of illness scoring; laboratory value (complete blood count with differentials, serum creatinine); SpO_2_:FiO_2_ ratio or arterial blood gases results (PaO_2_/FiO_2_ ratio, PaCO_2_, pH, O_2_ saturation); SpO2:FiO2 ratio will be recorded prior to proning, post proning at one hour, four hours and eight hours from initiation of proning; hemodynamic variables including MAP, HR, inotropic and vasoactive agents, intubation status, time to intubation, NIPPV settings, daily fluid balance, renal replacement therapy (RRT) use; sedative agents, corticosteroids use, antibiotics use, anticoagulation use, antiviral use and specific COVID-19 therapies, and survival status at 60 days. Daily data collection will continue until proning discontinuation criteria is met, even if the proning intervention was discontinued on the ward.

#### 4.2.2 Data Quality Monitoring

The Research Coordinator will enter data electronically through REDCap system. Quality control will include regular data verification and protocol compliance checks by the Study Coordinator. The Study Coordinator will also complete final reports detailing the recruitment, adverse events, and protocol deviation. The reports will be reviewed by the principal investigator and the members of the methods centre.

#### 4.2.3 Data Storage

The research team at each participating site will collect and record study data, as outlined in the data collection system REDCap. On all study documents, specific patient data will be identified with unique study identifiers. Trial data will be stored in a secured hospital network drive.

Once the trial recruitment period is complete, all study data will be securely stored in the Critical Care Hamilton storage facility for 15 years. At the end of the storage period, all study-related electronic stored information will be destroyed by a bonded company, according to institution privacy standards.

### 4.3 Safety Outcomes (Adverse events reporting)

An adverse event is any unfavourable and unintended sign, symptom, syndrome, or illness that develops or worsens during the period of the study. The overall risk of an adverse event occurring that is directly connected to this study procedure is likely to be minimal such as central or peripheral line dislodgment, fall, hypotension, pressure ulcers. To mitigate these possible adverse events, every effort will be made to secure the intravenous accesses; keep patients *nil per os* (NPO) in case the need arises for emergent intubation, based on treating team discretion. The patient will be monitored hourly in the ICU with frequent blood gases. If patients were to get worse, the treating team will take the necessary measure to intubate these patients in a timely manner as usual.

A Serious Adverse Event (SAE) is any adverse event occurring following study mandated procedures, directly related to the treatment or intervention that results in any of the following outcomes: Death, life-threatening adverse event, disability, or incapacity.

All intervention-related SAE will be recorded and reported to the institutional REB. Unexpected SAE will be reported within the mandated time frames to the institutional REB. SAEs will be defined and managed according to Clinical Safety Data Management Definitions and Standards for Expedited Reporting ICH Topic E2A (website: http://www.hc-sc.gc.ca/dhp-mps/prodpharma/applic-demande/guide-ld/ich/efficac/e2a-eng.php). All SAEs will be documented in detail on the appropriate case report form. During weekly method centre meetings, the SAE forms from the previous week will be reviewed and followed until satisfactory resolution. The Study Investigators will be immediately informed about any SAEs.

### 4.4 Participant withdrawal

Participants will be withdrawn from the study if they withdraw consent at any time with or without providing a reason, and if it is deemed unsafe, or is found to be impossible to proceed with any study procedure.

## 5.0 Ethical Considerations

This study was reviewed and approved by the REB prior to the initiation of the trial. Participation in this study is completely voluntary, and patients will suffer no penalty or loss of any benefits to which they would otherwise be entitled, should they decide not to participate. Any protocol amendment and/or deviations, and adverse events will be reported to the REB according to Health Canada regulatory and International Council for Harmonization-Good Clinical Practice (ICH GCP) standards. The trial will be conducted in compliance with the protocol, the ICH GCP Guidelines, the guiding principles of the Declaration of Helsinki, and all applicable regulatory requirements.

### 5.1 Confidentiality

Principles of privacy and confidentiality will be followed, whereby the identity and records of the subjects of the study will be kept confidential, and no details about the subjects that would result in the disclosure of their identities will be revealed. We are assuring that personal information provided by applicants will be protected, and at no stage will it form any part of the assessment process. Access to all patients’ data will be restricted to the domain of the research only based on the ethics approved data collection parameters.

## 6.0 Study Organization

### 6.1 Coordination and study management

Site initiation will be led by the Trial Manager along with the Primary Investigators and local Principal Investigator to ensure all the relevant paperwork and the standard operating procedure are applied at each participating centre. Regular methods centre meetings will be held weekly with primary investigators, Trial Manager, and trial coordinators. While quarterly meetings will be held between the primary investigators and the collaborators and site chiefs to assess the study progress, recruitment rate, and logistical issues need to be addressed. Monthly newsletter addressing relevant logistical study matters, recruitment rate, and study updates will be sent to each participating centre.

### 6.2 Steering committee

Dr. Alhazzani, the Principal Investigator, will lead regular meetings with the steering committee to assess study feasibility and progress, and any amendments to the study protocol. The steering committee includes multinational senior Canadian Institute of Health Research (CIHR)-funded trialists, senior biostatistician, and senior trial manager. The active Steering Committee members include Drs. Zainab Al Duhailib (Co-PI), Yaseen Arabi, Emilie Belley-Cote, Eddy Fan, Morten Hylander Moller, Dan Perri, Ken Parhar, Jason Weatherald, Bram Rochwerg, Muhammed Alshahrani, and Deborah Cook.

## 7.0 Knowledge translation

During the conduct of the study, brochures as well as presentations in grand rounds or conferences, will be held to the relevant healthcare workers to highlight the importance of conducting the study and answering this relevant question during the COVID-19 pandemic to ensure engaging each participating centre.

After the study conduct, the Principal Investigators and the Steering Committee will ensure publication of the study in a peer-reviewed journal. The results of the COVI-PRONE trial will be included in an individual-patient data meta-analysis with other similar trials. Lastly, the results of this trial will be incorporated into international clinical practice guidelines to ensure knowledge dissemination.

## 8.2 Trial Status

As of May 18 2021, 400 patients have been recruited, and participating sites are collecting 60-day outcome data.

The trial investigators and the trial statistician remain blinded to the outcome data, the protocol was submitted prior to completion of data collection and data cleaning. We anticipate completing the analysis and finalizing the first draft of the manuscript in September 2021.

## Data Availability

not applicable

## 9.0 Declarations

### 9.1 Ethics approval and consent to participate

The study was approved by Clinical Trials Ontario (CTO) ethics committee (reference number 2154), as well as the respective ethics boards of participating centres not governed by CTO, as per the list below. Eligible study participants will be enrolled and consent will be obtained through a mixed model using a priori or deferred consent as appropriate.

The list of regulatory bodies that have approved the study conduct:

1. Clinical Trials Ontario (Canada) -All Ontario Sites Clinical Trials Ontario Project ID 2154
2. The Conjoint Health Research Ethics Board (Canada) -All Calgary Sites REB 20-0731
3. State of Kuwait Ministry of Health, Assistant Undersecretary for Planning & Quality (Kuwait) -Both Sites Study No. 1526/2020
4. The Research Ethics Board (REB) of the Centre Hospitalier Universitaire de Québec, Université Laval (Canada) -Quebec MP-20-2021-5459
5. King Abdullah International Research Center (Saudi Arabia) -Sites 176, 177, 185, and 211 IRBC/0823/21
6. King Abdullah International Research Center (Saudi Arabia) -Site 185 IRBC/ 1181/20
7. Imam Abdulrahman Bin Faisal University Institutional Review Board (Saudi Arabia) -Site 178 IRB-2020-01-180
8. Committee of the Protection of Human Subjects (USA) -Texas HSC-MS-20-0793
9. King Faisal Specialist Hospital and Research Centre, Research Ethics Committee (Saudi Arabia) – Site 179 RAC 2201114

### 9.6 Authors’ contributions

WA and ZA conceived the idea, designed the study protocol, and wrote the first draft and subsequent drafts. All other authors contributed to study design, critical review of the protocol. HT contributed to the statistical analysis plan critical review.

## 9.7 Acknowledgements

We would like to acknowledge all the members of the steering committee, data and safety monitoring committee, and all our collaborators.

