## Supplement for "Awake Prone Position in Hypoxemic Patients with Coronavirus Disease 19 (COVI-PRONE): A Study protocol and Statistical Analysis Plan for Randomized Clinical Trial"

**Correspondence:**

Follow-Up	1

Enrollment	1

### 1 Abbreviations and Definitions

| AE | Adverse Event |
| --- | --- |
| COVID-19 | coronavirus infection 2019 |
| CI | Confidence interval |
| DSMC | Data safety and monitoring committee |
| GCS | Glasgow coma scale |
| HFNC | High flow nasal cannula |
| HR | Hazard ratio |
| ICU | Intensive care unit |
| NIPPV | Non-invasive positive pressure ventilation |
| RRR | Relative risk reduction |
| SOC | Standard of Care |
| SPO2:FiO2 | Ratio of peripheral oxygen saturation to fractional inspired oxygen |
| SAE | Serious adverse event |

### 2 Introduction

#### 2.1 Preface

Mortality of intubated patients with COVID-19 is high and innovative approaches to improve the outcomes of these patients has been sought by the scientific community. Awake prone position for hypoxic respiratory failure patients is simple and easy intervention. It has gain wide interest in light of the scarce resources and mechanical ventilators to assist in relieving hypoxia from recent evidence from observational trials, but the effect on other patient-important outcomes is uncertain. The aim of the COVI-PRONE Trial is to determine if early awake prone positioning in COVID-19 patients with hypoxemic respiratory failure; irrespective of the mode of oxygen delivery; reduces the need for invasive mechanical ventilation.

#### 2.2 Scope of the analyses

These analyses will assess the efficacy and safety of awake prone position versus standard of care (SOC), which is considered no prone positioning in non-intubated COVID-19 patients with hypoxemia on reducing the need for invasive mechanical ventilation. This analysis will be included in the final clinical study report.

### 3 Study Objectives and Endpoints

#### 3.1 Study Objectives

To determine the effect of early awake prone positioning versus SOC (no prone positioning) in non-intubated COVID-19 patients with hypoxemia on the need for invasive mechanical ventilation.

#### 3.2 Endpoints

#### 3.2.1. Primary Outcome

1. Endotracheal intubation within 30 days of randomization.

#### 3.2.2. Secondary Outcomes

1. Mortality at 60 days
2. Days alive and free of invasive mechanical ventilation at 30 days.
3. Days alive and free of non-invasive ventilation at 30 days
4. Days alive and out of the ICU (i.e., ICU free days), censored at 60 days.
5. Days alive and out of the hospital (i.e., Hospital free days), censored at 60 days.

**3.2.2.1. Safety Outcomes**

1. Complications from proning, which includes any of the following: accidental removal of intravenous access, hypotension, falls from bed, pressure ulcers, or other adverse events as defined below.

- **a) An adverse event (AE) is defined as:**

Any adverse event occurring following study mandated procedures, directly related to the treatment or intervention that results in any of the following outcomes: removal of intravenous access, hypotension, falls, pressure ulcers or other complications.

- **b) A serious adverse event (SAE)* in the COVI-PRONE trial is defined as:**

a) Any adverse event occurring following study mandated procedures, directly related to the treatment or intervention that results in any of the following outcomes: Death, life-threatening, adverse event, disability, or incapacity. **OR**

b) Any event that may jeopardize the patient and requires medical or surgical intervention to prevent one of the outcomes listed above. **AND**

c) Which the attending physician perceives may be directly related to enrolment in the COVI-PRONE trial

*Since ICU patients commonly develop complications of critical illness, related or unrelated to the reason for their admission to ICU (e.g., nosocomial infection, organ failure, and myocardial infarction), these often-expected events in the course of patients requiring life support will not be reported as SAEs.

### 4 Study Methods

#### 4.1 General Study Design and Plan

- **Study configuration and experimental design:** multicenter parallel group stratified pragmatic randomized controlled trial
- **Type of Comparison:** superiority.
- **Type of control(s):** SOC (i.e., no prone positioning).
- **Level and method of blinding:** Data collectors and healthcare team will be unblinded to the study arms trial due to the nature of prone positioning, however, the investigators will be blinded to outcome data until the final analysis is complete.
- **Method of treatment assignment:** Randomization is be based on variable undisclosed block sizes (2, 4, and 6 blocks) using a computer-generated table on ([www.Randmoize.net](https://randomize.net/)), randomization will be stratified based on hypoxia severity and recruiting center.
- Patients are randomized as soon as they meet the inclusion criteria and agree to participate in the trial.
- **Sequence and duration of all study periods:** patients will be screened for inclusion and exclusion criteria and if eligible they will be enrolled. Patients will be Randomized to either the intervention (prone position) or the control arm (SOC – no prone positioning). Daily data collection will continue until meeting one of the stopping criteria. We will follow the patient up to 60 days for the mortality outcome and for the days alive and out of the ICU and days alive and out of hospital.

#### 4.2 Inclusion-Exclusion Criteria and General Study Population

**6.2.1. Inclusion Criteria**

1. Adults ≥ 18 years of age.
2. Suspected or confirmed COVID-19. Defined as: a positive PCR for SARS CoV-2 or pending PCR results for patients that are suspected to have COVID-19.
3. Hypoxemia that requires ≥ 0.4 FiO2 (i.e. ≥ 40% oxygen) or ≥ 5L/min nasal cannula.
4. Bilateral or unilateral chest infiltrates on x-ray as interpreted by the treating team.
5. Admitted to the ICU or an acute care unit where hemodynamic and respiratory monitoring is feasible.

**6.2.2. Exclusion Criteria**

1. Immediate need for intubation as determined by the treating team.
2. Decreased level of consciousness (Glasgow Coma Scale score <10), or significant cognitive impairment that may interfere with compliance (delirium, dementia)
3. Contraindication to proning, including but not limited to any of the following:
   a. Open chest or abdomen, abdominal surgery (i.e. laparotomy) within the 4 days,
   b. Unstable spine, facial, cervical, femur, or pelvic fractures.
   c. Limited neck mobility or inability to lie prone comfortably
   d. Skeletal deformities that interfere with proning.
   e. Complete bowel obstruction.
   f. Active upper gastrointestinal bleeding.
   g. Patient is unlikely/unable to prone, or to be compliant as indicated by the treating team.
4. Body mass index > 40 kg/m2.
5. Pregnancy in the third trimester
6. Patient/substitute decision maker (SDM) or caring physician’s refusal to enroll in the study.
7. Patients with hemodynamic instability and moderate to high dose of vasopressors (norepinephrine dose ≥ 0.15 mcg/kg/min or equivalent)
8. Intubation is not within patient`s goals of care
9. Patient received awake proning session for > 1 day prior to randomization
10. Previous intubation within the same hospital admission

**6.2.3. General population**

Adult COVID-19 patients in the ICU or acute care unit who are receiving oxygen therapy for hypoxia, irrespective of the oxygenation modality

#### 4.3 Randomization and Blinding

**6.3.1. Randomization**

Randomization of study participants will be based on variable undisclosed block sizes (2, 4, and 6 blocks) through a computer-generated table using Randmoize.Net (http://www.randomize.net/). The randomization algorithm prepared by a biostatistician will stratify patients by the site of recruitment and severity of hypoxemia. Severity of hypoxemia will be classified based on the ratio of peripheral oxygen saturation to fractional inspired oxygen (SpO2:FiO2) into two strata: SpO2:FiO2 > 150 and SpO2:FiO2 ≤ 150 immediately prior to randomization. We estimated the SpO2:FiO2 thresholds using the following equation: SpO2/FiO2 ratio = 68 + 0.84 x (PaO2/FiO2 ratio) (21). Study participants will be randomized to either the intervention arm (awake prone positioning) (nasal cannula or oxygen mask or HFNC or NIPPV with prone positioning) or the control arm(SOC – no prone positioning). Concealment of randomization will be ensured at each center through a remote dedicated randomization system.

**6.3.2. Blinding**

Blinding of bedside healthcare workers and data collectors is not possible due to the nature of the prone positioning procedure. It is not possible to blind the trial biostatistician as the two groups will have clear separation in hours of proning which will preclude blinding to outcome data. However, study investigators and steering committee members will be blinded to outcome data until final analysis is complete.

#### 4.4 Study Assessments

**Figure S1.**

Eligible hypoxemic COVID-19 patient

Prone position

No prone position

Randomization

Stopping criteria

1. 40% RRR in FiO2 from baseline for 24 hours

2. Intubation

3. Discharge from ICU

4. Death

Daily Data collection

Complication of proning

30-day follow up

1. Invasive mechanical ventilation
2. Days alive and free of invasive mechanical ventilation
3. Days alive and free of non-invasive ventilation
4. Complication from proning

60-day follow up

1. Mortality

2. Days alive and outside the ICU

3. Days alive and outside the hospital

**Table S1. SpO2:FiO2 daily data collection**

| Intervention | Baseline | 1 hour | 4 hours | 8 hours | Daily AM |
| --- | --- | --- | --- | --- | --- |
| SpO2:FiO2 for Prone position | x | x | x | x | x |
| SpO2:FiO2 for SOC no prone positioning | x |  |  |  | x |

**Table S2. Daily data collection**

| Both groups | Baseline | Daily |
| --- | --- | --- |
| Mode of oxygenation | x | x |
| S/F ratio | x | x |
| Proning hours |  | x |
| Co-interventions | x | x |
| Stopping criteria |  | x |
| Adverse events |  | x |

### 5 Sample Size

We originally estimated the study sample size to be 350 patients. This was based on a relative risk reduction (RRR) of 25% (1), a 60% intubation rate in the control arm (2), and a power of 80%, and two-sided type I error of 5%. As the management of COVID-19 evolved and recent guidelines suggesting a trial of HFNO or NIPPV prior to intubation, the threshold for intubation of COVID-19 patients changed. Therefore, we increased the sample size by 14% (i.e. target sample size of 400 participants) which allows 80% power to detect 30% RRR in the primary outcome given a control group event rate of 45%. This change was approved by the trial’s Steering Committee, the Data Safety and Monitoring Committee (DSMC), and research ethics board (REB).

### 6 General Analysis Considerations

#### 6.1 Timing of Analyses

The final analysis will be performed after this SAP document is finalized, the last alive patient has completed the 60 day follow-up, the dataset is complete and locked. At that time, the cleaned dataset will be transferred to statistician for analysis.

#### 6.2 Analysis Populations

- **The intervention group:** subjects who were randomized to the proning arm.
- **The control group:** subjects who were randomized to the SOC no prone positioning arm.
- **The primary efficacy endpoint:** rates of intubation at 30 days of randomization.
- **The secondary endpoint:**
  - **Efficacy endpoints:** mortality at 60 days from randomization, invasive and non-invasive ventilation free days at 30 days, ICU free days at 60 days, and hospital free days at 60 days
  - **Safety endpoint:** central or peripheral line dislodgment, vomiting, falls, and pressure ulcers. Serious adverse events: any adverse event occurring following study mandated procedures, directly related to the treatment or intervention that results in any of the following outcomes: death, life-threatening adverse event, disability, or incapacity.

##### 6.2.1 Full Analysis Population (Intention to Treat)

- All subjects who were randomized and had complete outcome data.

##### 6.2.2 Safety Population

- All subjects who received any study intervention (including control) and are confirmed as providing complete follow-up regarding adverse event information but excluding subjects who were allocated to the intervention arm but did not receive the intervention.

#### 6.3 Covariates and Subgroups

We planned *a priori* 5 subgroup analyses for the primary outcome based on:

1) Severity of hypoxemia at randomization classifying subgroups into SPO2:FiO2 ratio > 150 versus ≤ 150 (we hypothesize that the treatment effect will be larger in patients with severe hypoxia)

2) Disease status classifying subgroups into SARS CoV-2 positive patients versus patients with negative testing for SARS CoV-2 (assuming that treatment effect will be larger in the SARS CoV-2 positive subgroup)

3) Initiated on nasal cannula or oxygen mask versus NIPPV versus HFNC (assuming that HFNC group will have a larger treatment effect)

4) Age ≥70 or <70 years old.

5) Sex

We will use test of interaction to compare subgroups. Results will be reported using RR and 95% CI and the multivariable logistic regression will be used to report the results of tests of interactions.

To assess the robustness of the results, we will perform the following *a priori* sensitivity analyses for the primary trial outcome:

1) Death as a competing risk

2) Per-protocol analysis (effectiveness analysis)

3) Per-site analysis.

##### 6.3.1 Multi-center Data

The data from multiple centers will be combined and analyzed as a whole. A sensitivity analysis will be conducted to test the center effect on outcomes.

#### 6.4 Missing Data

We will quantify the missing data and will report the reason for the missing data if applicable. We will not use multiple imputations to account for missing data.

#### 6.5 Interim Analyses and Data Monitoring

An independent DSMC will review the accumulating study data after 50% of patients have been randomized and make recommendations to the study leadership about the conduct of the trial, integrity of the data and trial discontinuation to ensure the overall safety of participants. The guiding policies and operating procedures governing the DSMC will be described in a separate DSMC charter.

The DSMC will follow a modified Haybittle-Peto boundary of 3 SD for benefit and harm as a general guideline. No modification of the level of significance of the results is necessary with the higher 3 SD boundary for the Haybittle-Peto criteria. The DSMC will also be monitoring the safety of the participants, focusing on adverse events and other safety indicators.

#### 6.6 Purpose of Interim Analyses

The purpose of the interim analysis is to ensure the appropriate conduct of the trial, integrity of the data, and the overall safety of participants and monitoring adverse events and to consider trial discontinuation if any safety concerns is raised by the DSMC.

The DSMC will hold one formal interim analysis meetings to monitor protocol adherence, intervention efficacy, participant safety, and quality of trial conduct.

The DSMC will be provided with a report from the biostatistician, which will include analysis of the primary outcome (rates of intubations), rates of serious adverse events, and rates of protocol violation. They will be provided with the recruitment rate and baseline characteristics of patients enrolled in both groups.

#### 6.7 Planned Schedule of Interim Analyses

##### The members of the DSMC will review the accumulating study data for the interim analysis when 50% of patients have been randomized.

##### 6.7.1 Scope of Adaptations

##### After the interim analysis, the DSMC will issue recommendation to enforce protocol adherence and a recommendation to stop or continue the trial will be issued from the DSMC.

##### 6.7.2 Stopping Rules

The DSMC will follow a modified Haybittle-Peto boundary of 3 SD for benefit and harm as a general guideline. No modification of the level of significance of the final results is necessary with the higher 3 SD boundary for the Haybittle-Peto criteria.

##### 6.7.3 Practical Measures to Minimize Bias

The data will be sent through secure transfer to the biostatistician to perform the interim analysis without any patient identifier. The generated report will be presented by the biostatistician who performed the data analysis to the DSMC members in the closed meeting. The DSMC will be unblinded to the treatment allocation due to the nature of the proning intervention.

The final analysis will be done by another independent biostatistician to reduce any bias from prior knowledge of the interim analysis results.

##### 6.7.4 Documentation of Interim Analyses

Records of the data available at the interim analysis should be preserved, as should all documentation of analysis plans, programming code and reporting will be provided at each interim.

#### 6.8 Multiple Testing

For secondary and safety outcomes, the False Discovery Rate will be calculated to account for multiple testing, which reflects on the expected proportion false positive tests.

### 7  Summary of Study Data

#### 7.1 Subject Disposition

A consort flow diagram will display the number of subjects screened, randomized and enrolled, withdrew consent, reached the final stages of the trial.

#### 7.2 Protocol Deviations

- A protocol deviation in the control arm is defined as the number of days with any proning hours before meeting the stopping criteria.
- A protocol deviation in the intervention arm is defined as zero proning hours across all days before meeting the stopping criteria.

Protocol deviations will be acknowledged and identified in the final analysis and publication manuscript. Patients will be analyzed in their respective group as they were originally allocated, using the intention-to-treat principle.

#### 7.3 Demographic and Baseline Variables

We will present patients’ demographics, clinical characteristics, and adverse events related to prone position as frequencies and percentages for categorical data (co-morbidities, sedation, vasoactive medication, antibiotics, anticoagulation, use of renal replacement therapy, and COVID-19 directed therapies, patients receiving nasal cannula/ face mask/ HFNC/ NIPPV/ invasive mechanical ventilation) and mean and standard deviation (SD) or median and interquartile range (IQR) for continuous data (vital signs, fluid balance, duration of proning in hours, number of breaks in hours from the start till the end of prone position, number of days in prone position, number of days with protocol deviation in both groups as defined above, FiO2 and SpO2 measurements prior and after prone position at one, four, and eight hours, mechanical ventilation parameters, and blood gases variables), as appropriate. Moreover, comparison between the 2 arms will be carried out using the chi-square or Fisher exact test for categorical variables, whereas the student’s t-test or Mann Whitney test will be used for the continuous ones.

#### 7.4 Treatment Compliance

Daily monitoring for compliance to the intervention or the control arm will be done through daily data collection sheets at the bedside and through documented data in the electronic medical records and paper charts. Health care teams are encouraged to ensure the patients compliance to the allocated treatment arm.

The proportions of patients receiving zero proning across the study period in the intervention arm will be reported. The proportion of patient’s receiving any proning in the control arm will also be reported.

### 8 Efficacy Analyses

- **The null hypothesis:** early proning of hypoxemic COVID-19 patients while receiving oxygen therapy, irrespective of the modality, does not reduce the need for invasive mechanical ventilation compared to no proning.
- **The alternative hypothesis:** early proning of hypoxemic COVID-19 patients while receiving oxygen therapy, irrespective of the modality, reduces the need for invasive mechanical ventilation compared to no proning.

All analyses will be performed using the Statistical Analysis Software (SAS version 9.4) (Cary, NC). Throughout the study, statistical significance will be set at alpha = 0.05.

#### 8.1 Primary Efficacy Analysis

We will use the intention-to-treat principle for the analysis of the data, in which trial participants will be analyzed in the group to which they were randomized.

The main analysis will be an unadjusted survival analysis (time-to-event) for the binary outcomes such as intubation, mortality at 60-day, considering the 2 stratified randomized variables of severity of hypoxemia and center. Comparison between the 2 arms will be carried out using the log-rank test for the time-to-event analyses. In this analysis we will use all information up to the time of censoring. We will calculate hazard ratios (HR) and associated 95% confidence intervals (CI) using Cox proportional regression analysis.

For continuous outcomes (i.e., hospital length of stay and ventilator free days), we will compare the 2 arms using a nonparametric approach to compare median durations because the distributions are likely skewed. Continuous outcomes will be analyzed using multiple linear regression, where our main independent variable will be the randomized group, and we will also include recruitment site and severity of hypoxemia as covariates. For this analysis, we will report the beta coefficient and the 95% CI.

#### 8.2  Secondary Efficacy Analyses

##### 8.2.1 Secondary Analyses of Primary Efficacy Endpoint

An efficacy analysis will be conducted including all patients who have undergone at least 8 hours of prone positioning daily until meeting discontinuation criteria (in the intervention arm) and those who did not undergo any hours of prone positioning (in the control arm).

##### 8.2.2 Analyses of Secondary Endpoints

See section 10.1

### 9 Safety Analyses

#### We will calculate the frequency of events post-randomization for each group and compare via a Pearson chi-square test. We will calculate the RR and corresponding 95% CI for this event.

| Adverse events | Prone Positioning | SOC  (no prone positioning) |
| --- | --- | --- |
| Accidental removal of intravenous access |  |  |
| Hypotension |  |  |
| Falls |  |  |
| Pressure ulcers |  |  |
| Other |  |  |

### 10 Reporting Conventions

P-values ≥0.001 will be reported to 3 decimal places; p-values less than 0.001 will be reported as “<0.001”. The mean, standard deviation, and any other statistics other than quantiles, will be reported to two decimal place greater than the original data. Quantiles, such as median, or minimum and maximum will use the same number of decimal places as the original data.

### 11 Summary of Changes to the Protocol

| Amendment Number & Date: | Summary of Changes: | IRB approval date: |
| --- | --- | --- |
| Amendment #1  Protocol V10.0  15-MAY-2020 | 1. **Modification to the inclusion criteria;**   Criteria #3: Removal of room air and SPO2 <90% and addition of or >5L nasal cannula.   2. **Addition of exclusion criteria;**   Criteria #8: Pregnancy in the third trimester  Criteria #9: Limited neck mobility or inability to lie prone comfortably   3. **Modification to the secondary Outcomes;**   Outcome #1: Mortality changed to 60 days  Outcome #6: Removal of vomiting from complications from proning   4. **Addition of secondary outcomes;**   Outcome #4: Days alive and out of the ICU, censored at 60 days.  Outcome #5: Days alive and out of the hospital, censored at 60 days   5. **Removal of secondary outcomes;**   Outcome #4: ICU length of stay censored at 30 days  Outcome #5: Hospital length of stay censored at 30 days  Outcome #6: Change in oxygen defined as the difference in SPO2:FiO2 ratio at 1 hour, 4 hours, and 8 hours.   6. **Modification to the proning discontinuing criteria**;   SPO2:FiO2 ratio > 300 and FiO2 <0.3 (ie. ≤ 2 L nasal cannula) for 24 hours.   7. **Modification to the Subgroup Analysis;**   Subgroup #1: SPO2:FiO2 ratio stratification defined: >150 versus ≤150.   8. **Addition to the Subgroup Analysis;**   Subgroup #4: Age ≥ 70 or <70 years old.  Subgroup #5: Sex | 19-MAY-2020 |
| Amendment # 2  Protocol V11.0  31-JUL-2020 | 1. **Additional Exclusion criteria;**   Criteria #11: Patient received awake proning session for >1 day prior to randomization.  Criteria #12: Previous intubation within the same hospital admission.   2. **Modification to the Proning Discontinuation Criteria;**   Criteria #1: Changed to: relative reduction improvement in FiO2 of 40% from the baseline FiO2 value for 24 hours.  Criteria #3: Added: discharge from the ICU or the acute care monitoring unit, in which patient’s oxygenation cannot be monitored.   3. **Data Collection Modification;**   Addition of protocolized data collection for patients discharged from acute care setting prior to stopping criteria, to be followed until a stopping criterion has been met. | 12-AUG-2020 |
| Amendment #3  Protocol V12.0  03-OCT-2020 | 1. **Modification and reformatting to the exclusion criteria;**   Criteria #2 Specification added: Decrease level of consciousness (Glasgow Coma Scale score <10)  Criteria #3 Contraindications to proning consolidated and elaborated to include:   - Unstable spine, facial, cervical, femur, or pelvic fractures - Limited neck mobility or inability to lie prone comfortably - Skeletal deformities that interfere with proning. - Complete bowl obstruction - Active upper gastrointestinal bleeding. - Poor neck mobility or patient inability to lie prone comfortably.   Criteria # 4: Body mass index > 40 kg/m2 made a stand-alone | 02-NOV-2020 |
| Amendment #4  Protocol V13.0  09-NOV-2021 | 1. **Removal of repeating exclusion criteria;** | 28-NOV-2020 |
| Amendment #5  Protocol V14.0  25-JAN-2021 | 1. **Modification to the exclusion criteria;**   Criteria #2 elaborated to include: significant cognitive impairment that may interfere with compliance (delirium, dementia).  Criteria #3 elaborated to include:   - Patient is unlikely/unable to prone, or to be compliant as indicated by the treating team. | 04-FEB-2021 |
| Amendment #6  Protocol V15.0  09-APR-2021 | 1. **Increase in trial sample size;**   From 350 to 400 patients | 13-APR-2021 |

### 13  Tables and Figures

**1. Study Flow Diagram**

Assessed for eligibility (n= )

Excluded (n=	)

♦  Not ‑meeting inclusion criteria (n= )

♦  Declined to participate (n= )

♦  Other reasons (n= )

Analysed (n= )
♦ Excluded from analysis (give reasons) (n= )

Lost to follow-up (give reasons) (n= )

Discontinued intervention (give reasons) (n= )

Allocated to intervention (n= )

♦ Received allocated intervention (n= )

♦ Did not receive allocated intervention (give reasons) (n= )

Lost to follow-up (give reasons) (n= )

Discontinued intervention (give reasons) (n= )

Allocated to intervention (n= )

♦ Received allocated intervention (n= )

♦ Did not receive allocated intervention (give reasons) (n= )

Analysed (n= )
♦ Excluded from analysis (give reasons) (n= )

#### Allocation

#### Analysis

#### Follow-Up

Randomized (n= )

#### Enrollment

**Table 1. Patient’s baseline characteristics**

| **Characteristic** | **Prone positioning**  **(N)** | **Control**  **(N)** |
| --- | --- | --- |
| **Age — yr** | Median years (IQR) | Median years (IQR) |
| **Male sex — no. (%)** | N (%) | N (%) |
| **Race or ethnic group — no./total no. (%)**  Asian  Black  Mixed/Multiple  Native American  Native Hawaiian  White  Other  Middle Eastern | N (%)  N (%)  N (%)  N (%) N (%)  N (%)  N (%)  N (%) | N (%)  N (%)  N (%)  N (%) N (%)  N (%)  N (%)  N (%) |
| **Body-mass index** | Median Kg/m^2^ (IQR) | Median Kg/m^2^ (IQR) |
| **GCS** | Median (IQR) | Median (IQR) |
| **Median days to enrollment (IQR)**  **(time from hospital admission to time of randomization)** | Median days +/- SD | Median days +/- SD |
| **Confirmed SARS‑CoV‑2 infection — no./total no. (%)**  COVID Positive  COVID Negative | N (%) | N (%) |
| **SpO2/FiO2 at randomization** | Median (IQR) | Median (IQR) |
| **Inspired Oxygen (%)** | Median % (IQR) | Median % (IQR) |
| **Severity of hypoxemia**  SpO2/FiO2> 150  SpO2/FiO2≤ 150 | N (%)  N (%) | N (%)  N (%) |
| **Oxygen delivery mode**  NIPPV  HFNC  Low Flow | N (%)  N (%)  N (%) | N (%)  N (%)  N (%) |
| **Comorbidities**  Coronary Artery Disease  Congestive Heart Failure  Hypertension  COPD  Asthma  Respiratory others  VTE  Cirrhosis  Diabetes  Immunocompromised  Renal other  ESRD  Malignancy  Transplant | N (%)  N (%)  N (%)  N (%)  N (%)  N (%)  N (%)  N (%)  N (%)  N (%)  N (%)  N (%)  N (%) N (%) | N (%)  N (%)  N (%)  N (%)  N (%)  N (%)  N (%)  N (%)  N (%)  N (%)  N (%)  N (%) N (%)  N (%) |
| **Drugs**  Current Pre-Hospital Meds (check box)  Ace inhibitors  Angiotensin 2 receptor blockers  In-Hospital Meds – Pre Rand  Inotropic Agents  Vasoactive Agents  Sedatives  Corticosteroids  Methylprednisone  Hydrocortisone  Prednisone  Dexamethasone  Other  Antibiotics  Anticoagulation | N (%)  N (%)  N (%)  N (%)  N (%)  N (%)  N (%)  N (%)  N (%)  N (%)  N (%)  N (%)  N (%) | N (%)  N (%)  N (%)  N (%)  N (%)  N (%)  N (%)  N (%)  N (%)  N (%)  N (%)  N (%)  N (%) |
| **Anticoagulation Dose**  Therapeutic  Prophylactic | N (%)  N (%) | N (%)  N (%) |

**Table 2.**

| **Proning** | **Prone positioning**  **(N)** | **Control**  **(N)** | **Relative effect**  **(95% CI)** |
| --- | --- | --- | --- |
| Hours of proning per day  Prone Position  Control | Median hours (IQR) | Median hours (IQR) | Difference (95% CI) |
| Number of days in prone while meeting criteria  Prone Position  Control | Median days (IQR) | Median days (IQR) | Difference (95% CI) |
| Co-Interventions (Post Rand)  Oxygenation Mode  NIPPV  HFNC  Low Flow  RRT  Fluid Balance  Steroids  Methylprednisone  Hydrocortisone  Prednisone  Dexamethasone  Other  COVID-19 SOC  COVID Treatments  Hydroxychloroquine or Chloroquine  Hydroxychloroquine or Chloroquine + Azithromycin  Kaletra (lopinavir/ ritonavir)  Remdesivir  Convalescent plasma  Tocilizumab  Favipiravir  Interferon  Other  Anticoagulation  Dalteparin  Enoxaparin  Heparin  Tinzaprin  Apixaban  Rivaroxaban  Dabigatran  Warfarin  Other | N (%)  N (%)  N (%)  N (%)  Median days (IQR)  N (%)  N (%)  N (%)  N (%)  N (%)  N (%)  N (%)  N (%)  N (%)  N (%)  N (%)  N (%)  N (%)  N (%)  N (%)  N (%)  N (%)  N (%)  N (%)  N (%)  N (%)  N (%) N (%) | N (%)  N (%)  N (%)  N (%)  Median days (IQR)  N (%)  N (%)  N (%)  N (%)  N (%)  N (%)  N (%)  N (%)  N (%)  N (%)  N (%)  N (%)  N (%)  N (%)  N (%)  N (%)  N (%)  N (%)  N (%)  N (%)  N (%)  N (%)  N (%) |  |
| **Anticoagulation Dose**  Therapeutic  Prophylactic | N (%)  N (%) | N (%)  N (%) |  |

**Table 3. Primary outcome**

| **Outcome** | **Prone positioning**  **(N)** | **Control**  **(N)** | **Relative effect**  **(95% CI)** |
| --- | --- | --- | --- |
| **Primary Outcome** | | | |
| Endotracheal intubation at 30 days | N (%) | N (%) | HR (95% CI) |
| **Secondary Outcomes** | | | |
| Mortality at 60 days | N (%) | N (%) | RR (95% CI) |
| Days alive and free of mechanical ventilation at 30 days  Days alive and free of non-invasive ventilation at 30 days | Median (IQR)  Median (IQR) | Median (IQR)  Median (IQR) | Difference (95% CI)  Difference (95% CI) |
| Days alive and outside of ICU at 60-day | Median (IQR) | Median (IQR) | Difference (95% CI) |
| Days alive and outside of the hospital at 60-day | Median (IQR) | Median (IQR) | Difference (95% CI) |
| Safety Outcomes  Removal of IV access  Hypotension  Fall  Pressure ulcers  Other  AE/SAE  AE  SAE | N (%)  N (%)  N (%)  N (%)  N (%)  N (%)  N (%) | N (%)  N (%)  N (%)  N (%)  N (%)  N (%)  N (%) |  |

**Table 4. Protocol deviations**

|  | Prone^*^  *n =* | SOC^**^  *n =* |
| --- | --- | --- |
| Protocol Deviation: *n (%)* |  |  |

*Prone Group: Zero hours of prone position (across all days up to meeting stopping criteria – 40% relative reduction in FiO2, intubation, discharge, death)
**SOC Group: Any hours of prone position > zero (across all days up to meeting stopping criteria – 40% relative reduction in FiO2, intubation, discharge, death)

**Figure 2. Number of proning hours per group**

Number of hours spent in prone position

|  | Number of prone position hours |
| --- | --- |
| 1 | 1 |
| 2 | 2 |
| 3 | 3 |
| 4 | 4 |
| 6 | 5 |
| 7 | 6 |
| 8 | 7 |
|  | 8 |
|  | 9 |
|  | 10 |

**Figure 3. Oxygenation - SF Ratio in the first 7 days**

**Prone vs soc daily SF ratio**

SF Ratio

|  | Number of prone position hours |
| --- | --- |
| 1 | 1 |
| 2 | 2 |
| 3 | 3 |
| 4 | 4 |
| 6 | 5 |
| 7 | 6 |
| 8 | 7 |
|  | 8 |
|  | 9 |
|  | 10 |

**Figure 4. Prone change in SF ratio (Pre, 1hr, 4hr, 8hr)**

SF Ratio

|  | Number of prone position hours |
| --- | --- |
| 1 | 1 |
| 2 | 2 |
| 3 | 3 |
| 4 | 4 |
| 6 | 5 |
| 7 | 6 |
| 8 | 7 |
|  | 8 |
|  | 9 |
|  | 10 |

**Figure 5. Kaplan-Meier curve for the primary outcome (intubation)**

|  | No proning | Prone position |
| --- | --- | --- |
| 1 | 4 | 2 |
| 2 | 4 | 2 |
| 3 | 6 | 3 |
| 4 | 10 | 5 |
| 5 | 20 | 10 |
| 6 | 40 | 20 |

**Figure 6. Subgroup analysis – Forrest plot of subgroups**
